# A multi-centre prospective study of COVID-19 transmission following outpatient Gastrointestinal Endoscopy in the United Kingdom

**DOI:** 10.1101/2020.08.02.20166736

**Authors:** Bu’Hussain Hayee, The SCOTS project group, James E. East, Colin Rees, Ian Penman

**Affiliations:** King’s Institute of Therapeutic Endoscopy, King’s College Hospital NHS Foundation Trust, London, UK; Translational Gastroenterology Unit, Nuffield Department of Medicine, John Radcliffe Hospital, University of Oxford, Oxford, UK; Oxford NIHR Biomedical Research Centre, University of Oxford, Oxford, UK; Population Health Sciences Institute, Newcastle University, UK; Centre for Liver & Digestive Disorders, Royal Infirmary of Edinburgh, Edinburgh, UK

**Keywords:** COVID-19, Personal Protective Equipment, Gastroenterology, severe acute respiratory syndrome coronavirus 2, Endoscopy, Outpatient, Swab, Telephone, Aerosol

## Abstract

**Message:** The COVID-19 pandemic has severely curtailed the practice of endoscopy (as an exemplar for outpatient diagnostic procedures) worldwide. Restart and recovery processes will be influenced by the need to protect patients and staff from disease transmission, but data on the risk of COVID-19 transmission after endoscopy are sparse. This is of particular importance in later pandemic phases when the risk of harm from delayed or missed significant diagnoses is likely to far outweigh the risk of infection. The British Society of Gastroenterology (BSG) guidance for restarting endoscopy included stratification of diagnostic procedures according to aerosol generation or assessment of infectious risk as well as pragmatic guidance on the use of personal protective equipment (PPE). We sought to document the risk of COVID-19 transmission after endoscopy in this “COVID-minimised” environment. Prospective data were collected from 18 UK centres for n=6208 procedures. Pre-endoscopy, 3/2611 (0.11% [95% CI: 0.00-0.33%]) asymptomatic patients tested positive for SARS-CoV-2 on nasopharyngeal swab. Based on follow-up telephone symptom screening of patients at 7 and 14 days, no cases of COVID-19 were detected by any centre after endoscopy in either patients or staff. While these data cannot determine the relative contribution of each component of a COVID-minimised pathway, they provide clear support for such an approach. The rational use of PPE and infection control policies should be continued and will aid planning for outpatient diagnostics in the COVID-19 recovery phase.

## In more detail

The COVID-19 pandemic has had an extraordinary impact upon delivery of gastrointestinal (GI) endoscopy, with an initial reduction to 12% of pre pandemic levels in the United Kingdom (UK)[1], In the deceleration and early recovery phases (up to end July *2020)*, this had risen to 42% of re pandemic levels[2], Recovery has been influenced by multiple factors including availability of staff, restrictions caused by longer room cleansing, physical distancing and the use of personal protective equipment (PPE) slowing lists. There are grave implications of this contraction in activity[3], with the delayed diagnosis of significant conditions like GI cancer or inflammatory bowel disease of particular concern[1,4].

The risk around inadvertent peri-procedure transmission of COVID-19 infection to both patients and staff is a primary concern[4,5], but is not well described. Early data from northern Italy described low rates of infection in patients and staff, even during the peak phase of the pandemic[6].

Upper GI endoscopy is widely accepted to be an aerosol-generating procedure (AGP), however the relevance of small-volume aerosols (i.e. 0.3 micron; which appear to predominate[7]) for virus transmission is unclear[8]. Infectious potential appears to be confined to particles 0.5 micron or larger[8], but this is a complex subject with a number of variables. There are also direct data to support the effectiveness of masks, including surgical face masks most widely used in endoscopy units, in preventing viral transmission[9,10]. COVID-19 infection rates have been demonstrated to be lower-than-expected in endoscopy staff (compared to other health-care workers (HCWs) even when so-called ‘high-risk’ personal protective equipment (PPE; particularly face masks) were not universally available or applied[6]. Whether lower GI endoscopy is an AGP is also important but has been pragmatically regarded as having low infectious potential as per British Society of Gastroenterology (BSG) guidance[11], while staff are still advised to use appropriate (stratified) PPE for all procedures.

Significant patient anxiety regarding the potential for contracting COVID-19 infection also exists and this has also been demonstrated to influence the ability to provide effective diagnostic services[12]. We therefore sought to study prospectively the number of patient infections following GI endoscopy from multiple centres across the UK through the peak, deceleration and early recovery phases of the pandemic. Taking into account the complexities of infection control, aerosols, infectivity and abrogation of risk by PPE, it was felt that the ultimate determinant of risk would be whether any COVID-19 cases were actually detected after endoscopy (in either patients or staff). The purpose of this study was to enable quantification of potential risk to patients and to inform endoscopy departments regarding the likelihood of transmitting infection to patients. These data could be used to help communicate with patients and staff as well as inform planning for future outbreaks.

This multi-centre prospective study collected data for consecutive outpatients attending for elective diagnostic or therapeutic endoscopy from 18 UK centres. No patient identifiable data was collected, no treatment decisions were affected and no identifiable data were used, analysed or transferred. As such ethical approval was deemed not to be required by the Research Governance committee at the lead author’s institution.

Centres were selected to reflect different sized units, tertiary and local, covering a wide range of demographics including those serving mixed socioeconomic populations and a mix of ethnicities. All centres prospectively completed an anonymised database of patients including procedure type, responses to pre-procedure SCOTS criteria, pre-procedure nasopharyngeal swab test result (if performed in that unit), source of referral and dates for all activities. The SCOTS criteria[5] comprise telephone screening questions around Symptoms, infectious Contacts, Occupational risk, Travel risk, Shielding status[13] and are recommended in BSG guidance[11]. These were developed as an update to a pre-existing screening tool (FTOCC; developed in Hong Kong during the first SARS outbreak and proposed for using during the COVID-19 pandemic[14]), to take into account considerations specific to COVID-19.

All centres conducted patient follow-up by telephone consultation at 7 and 14 days after the procedure to check for symptoms of COVID-19 If symptoms were reported that could be consistent with COVID-19[5], these patients were advised to undergo COVID-19 nasopharyngeal swab testing. Data were collected on any patients with positive COVID-19 swab undertaken for any reason in the 14 days post endoscopy. Where patients were COVID-19 swab tested pre-procedure and found to be positive these patients were excluded from the follow-up study.

Data were collected from 6208 patients undergoing endoscopy at 18 centres between 30^th^ April and 30^th^ June 2020 (mean±SD age 59.3 ± 15.4 years and n=2973 (48%) were female). The characteristics of the endoscopy units are shown in table 1. Follow up data on symptoms were collected up to 14^th^ July 2020. There were no cases of COVID-19 detected in the two weeks following endoscopy (0/6208, 95% CI 0.0-0.08% with continuity correction).

**Table 1.** Characteristics of units submitting data.

| Site | Centre type | Annual procedure count (pre-covid) | Cases submitted | Pre-endoscopy actions |  | Stratified use† of ... |  |
| --- | --- | --- | --- | --- | --- | --- | --- |
|  |  |  |  | NP Swab? | Mandatory self-isolation for patients (≥ 7 days)? | PPE | IPCPs |
| A | T | 20000 | 198 | Yes | Yes | Yes | Yes |
| B | T | 17000 | 111 | No | No | Yes | Yes |
| C | L | 20000 | 436 | No | No | Yes | Yes |
| D | L | 9800 | 365 | Yes | No* | No | No |
| E | T | 15000 | 374 | Yes | No | Yes | Yes |
| F | L | 7500 | 58 | Yes | No | No | No |
| G | L | 9800 | 206 | Yes | No | Yes | Yes |
| H | T | 17000 | 218 | Yes | No* | Yes | Yes |
| I | L | 15200 | 351 | Yes | No | Yes | Yes |
| J | L | 17400 | 175 | Yes | Yes | Yes | Yes |
| K | T | 15000 | 84 | Yes | No | No | No |
| L | T | 15600 | 950 | No | No | Yes | Yes |
| M | T | 20000 | 363 | Yes | No | Yes | Yes |
| N | T | 30525 | 219 | Yes | No | No | No |
| O | T | 15000 | 715 | No | No | No | No |
| P | L | 24000 | 672 | No | No | No | No |
| Q | T | 14000 | 660 | No | No | Yes | Yes |
| R | L | 20000 | 251 | Yes | No* | Yes | Yes |
| **Total** | 10T; 8L | 302825 | 6208 | 12Y | 16N | 12Y | 12Y |

*\* sites suggested, but did not mandate, self-isolation of patients for 7 days pre-procedure*
*† Stratified use refers to the differential use of 'low-risk' and 'high-risk' principles according to upper vs lower GI endoscopy and COVID19-confirmed vs COVID19-excluded patients (ie. a COVID-minimised pathway as explained in references 4, 5 and 11)*
T – tertiary; L – local; NP – nasopharyngeal; PPE – personal protective equipment; IPCPs – infection prevention and control policies

Figure 1 shows the procedures performed, and overall % of total. There was an approximate 40:60 split between upper and lower GI procedures (where combined OGD and colonoscopy counted as upper GI – given the AGP status of the former procedure and therefore potential for greater risk).

**Figure 1.**
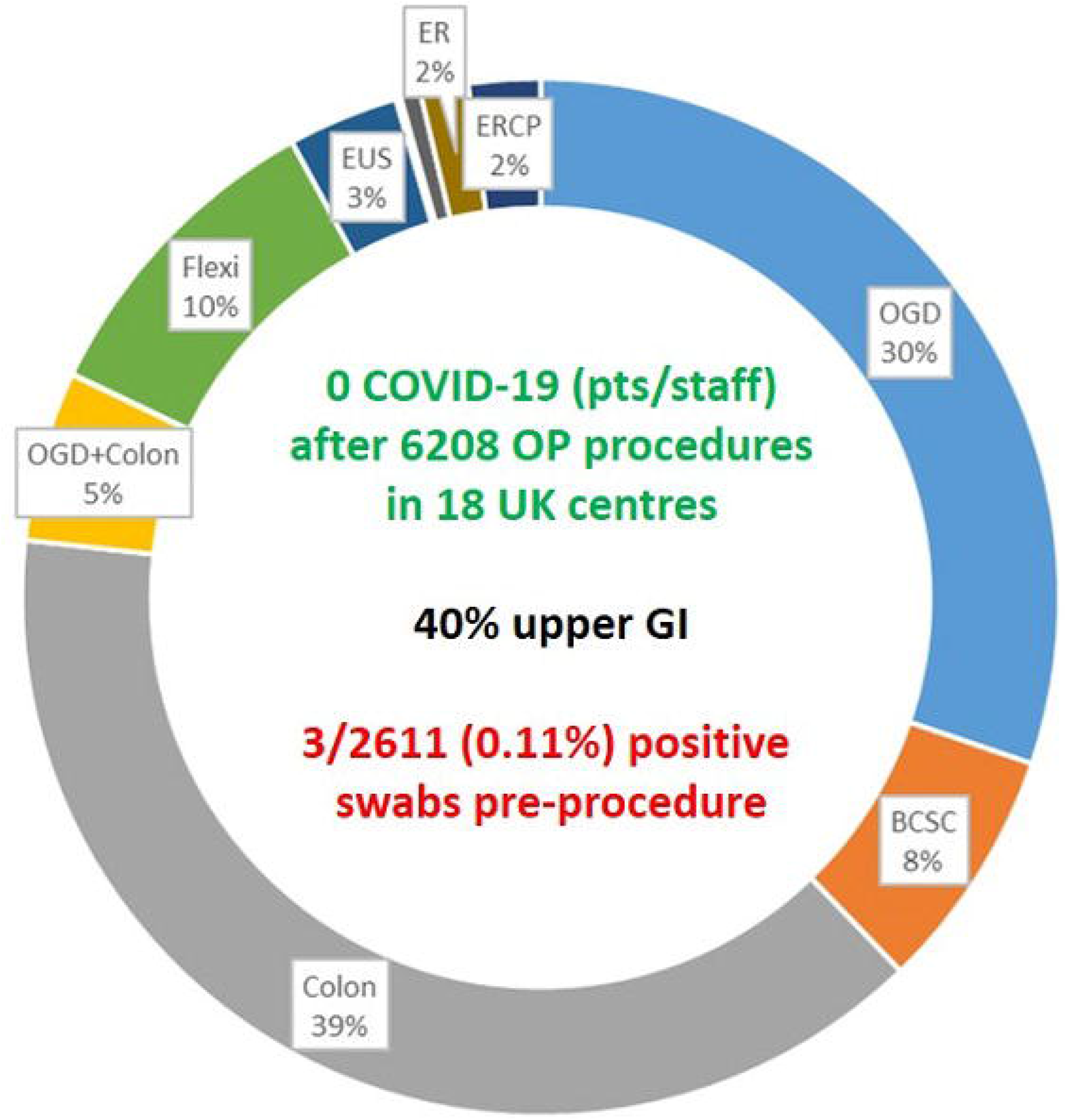
Proportions of procedures performed. Not specifically labelled on the figure are double balloon enteroscopy (n=1), percutaneous endoscopic gastrostomy (n=7), video capsule enteroscopy (n=28). Endoscopic resections (ER) were all lower GI. *OGD: oesophagogastroduodenoscopy; BCSC: bowel cancer screening colonoscopy (in national programme); EUS – endoscopic ultrasound; ER – endoscopic resection (endoscopic mucosal resection or endoscopic submucosal dissection); ERCP – endoscopy retrograde pancreatography; Flexi –flexible sigmoidoscopy; OP – outpatient*

Most centres were prioritising urgent symptomatic patients during this period with less than 4% (n=233) of procedures classed as “routine or surveillance” and these were performed at the discretion of the performing centre, mostly within the last two weeks of the data collection period. Of the remainder, n=3166 (51%), were patients referred for suspected cancer on the pre-existing UK “two-week wait”[15] pathways (again split 60:40 in favour of lower GI). There were 1193 (19.0%) in a ‘shielded’ category[13] as determined by pre-procedure telephone questionnaire.

Pre-procedure nasopharyngeal swab testing for SARS-CoV-2 was performed in 2611 patients from 13 sites, in line with BSG guidance, after confirmation of absence of COVID-19 symptoms or risks [5]. Only 3 (0.11%, 95% CI 0.03-0.36% with continuity correction) patients were positive. All had their investigation safely deferred with no complications.

Following their procedure 12 patients reported symptoms which were potentially compatible with COVID-19 infection at either the 7 or 14 day telephone contact. All then underwent nasopharyngeal swab testing and were found to be negative. All symptoms settled with none deemed to have COVID-19. There was, sadly, one reported death due to an entirely unrelated cerebrovascular event within 14 days of a colonoscopy.

## Comments

This prospective multi-centre study of 6208 patients provides a clear indication that GI endoscopy can be safely performed in the recovery phase of COVID-19, when a COVID-minimised pathway[5] is instituted. A relative strength of the study is that it involved 18 UK tertiary and local centres with a combined capacity of over 300,000 procedures per year.

There was a low incidence of positive swabs for asymptomatic patients pre-procedure, but telephone screening alone might have missed up to one in three hundred cases attending for endoscopy (at the top end of the 95% CI).

Our data cannot determine the relative contribution of each component of a COVID-minimised pathway (pre-procedure questionnaire, swab testing, use of PPE), but provides broad vindication of such an approach (a majority of the units involved used the recommendations for a stratified approach to PPE and infection control policies (air exchange, room cleaning, etc)). The combined effect of all these measures had a beneficial impact and should be continued through the pandemic. We cannot determine, in particular, whether less stringent PPE recommendations are possible beyond that already included in national guidance [11], but at the levels employed across the units involved in this study, there does not appear to be any excess risk.

New guidance in the UK, issued after the study period, now also recommends testing 48-72 hours preadmission for diagnostic tests requiring sedation[16], and this is likely a cost effective strategy (in a US context)[17]. This data collection was performed in the deceleration/recovery phases of the COVID-19 pandemic in the UK[18] and should therefore inform periods of activity where similar rates of infection are seen.

When a COVID-minimised pathway is in place, patients (including those in a high risk “shielding” category) can now be reassured that concerns over COVID-19 infection should not stop them attending for GI endoscopy.

## Data Availability

N/A

